# Determinants of cancer care delays in Kinshasa, Democratic Republic of the Congo (DRC)

**DOI:** 10.64898/2026.05.19.26353550

**Authors:** Jean Claude Dusingize, Natalia Zotova, Rafi Kabarriti, Kavita Sehrawat, Pélagie Babakazo, Ali Shongo Alisho, Fidele Lumande Kasindi, Ilyass Yessoufou, Marcel Yotebieng, CA-IeDEA

**Author notes:** Corresponding Author Jean Claude Dusingize, MD PhD Department of Medicine Albert Einstein College of Medicine.

## Abstract

**PURPOSE:** Cancer outcomes in sub-Saharan Africa are driven by delayed diagnosis and treatment initiation. We evaluated the magnitude and determinants of diagnostic and treatment delays among cancer patients in Kinshasa, Democratic Republic of the Congo (DRC).

**METHODS:** We conducted a hospital-based cross-sectional study of 460 adults with confirmed cancer at Nganda Hospital Center in Kinshasa, DRC. Two outcomes were assessed: delay from symptom onset to diagnosis and delay from diagnosis to treatment initiation. Log-normal regression models were fitted for each outcome to estimate adjusted geometric mean ratios (aGMRs) and 95% confidence intervals (CIs). Covariates included demographic, socioeconomic, clinical, behavioral, and stigma-related factors.

**RESULTS:** The median age was 55 years, and 76.2% of participants were women. Overall, 55.0% of participants experienced symptom-to-diagnosis delays >6 months, and 49.4% experienced diagnosis-to-treatment delays >3 months. Older age was associated with longer diagnostic delay (aGMR 1.55, 95% CI 1.03-2.31) and treatment delay (1.51, 1.07-2.14). Unemployment was strongly associated with both diagnostic delay (1.68, 1.15-2.47) and treatment delay (2.27, 1.54-3.33), as was hepatitis B co-infection (1.88, 1.06-3.34 and 2.42, 1.15-5.11, respectively). Longer diagnostic delay was additionally associated with informal trading (1.99, 1.21-3.28), taxi or motorbike transport (1.92, 1.25-2.94), and smoking history (2.25, 1.03-4.91), while high cancer-stereotype stigma was associated with longer treatment delay (1.56, 1.04-2.34).

**CONCLUSION:** Substantial delays exist across the DRC cancer care continuum, driven by socioeconomic vulnerability, transport barriers, hepatitis B co-infection, and cancer-related stigma. These findings highlight the need for integrated interventions to improve timely diagnosis and treatment initiation, including strengthening financial protection, decentralizing cancer services, and reducing stigma in cancer care.

## INTRODUCTION

The global burden of cancer is rising disproportionately in low- and middle-income countries (LMICs), which account for approximately 55-60% of new cancer cases and nearly 70% of cancer-related deaths worldwide (1,2). Survival remains substantially lower in these settings than in high-income countries, reflecting persistent inequities in access to timely diagnosis and treatment (3). Sub-Saharan Africa (SSA) is experiencing a rapid increase in cancer burden, compounded by the HIV epidemic and the high prevalence of oncogenic infections such as human papillomavirus (HPV) and hepatitis B virus (HBV), which elevates the risk of HIV-associated malignancies such as cervical cancer, Kaposi sarcoma, and non-Hodgkin lymphoma (4,5). A key driver of poor outcomes in the region is late-stage presentation, largely attributable to delays along the cancer care continuum from symptom recognition to diagnosis and from diagnosis to treatment initiation (6).

Delays in cancer diagnosis and treatment are multidimensional, stemming from a combination of patient-level factors (socioeconomic disadvantage, low health literacy, cultural beliefs and fear), provider-level factors (limited diagnostic capacity, inefficient referral pathways), and system-level challenges (geographic inaccessibility, high out-of-pocket costs) (7). In SSA, out-of-pocket expenditure on health is among the highest in the world as a share of total health spending, and cancer care is rarely covered by public programs, exposing patients to catastrophic financial risk (8). Cancer-related stigma, encompassing perceptions of fatalism, social discrimination, and stereotype-driven beliefs, has emerged as an underappreciated yet important sociocultural barrier to timely care-seeking in Low- and Middle-Income Countries (LMICs) contexts (9,10)

The Democratic Republic of the Congo (DRC) is one of the largest countries in sub-Saharan Africa, with a population exceeding 100 million and a gross domestic product (GDP) per capita of approximately USD 600 (11). The country faces a growing cancer burden in the context of constrained health system capacity and widespread poverty (12). Access to cancer care remains limited, with services concentrated in a small number of urban facilities and substantial out-of-pocket costs incurred by patients. Although recent national efforts aim to expand cancer services, including the establishment of the National Cancer Control Program (CNLC), there remains a critical need for data to inform strategies to improve timely access to diagnosis and treatment.

Despite substantial cancer care challenges in the DRC, quantitative data on delays in diagnosis and treatment initiation remain limited, and the role of cancer-related stigma has not been examined in this setting. We conducted a hospital-based cross-sectional study at Nganda Hospital Center, the country’s sole radiotherapy facility, to quantify delays across the cancer care continuum and identify their sociodemographic, occupational, clinical, and stigma-related determinants.

## METHODS

### Study design and population

We conducted a hospital-based cross-sectional study of adult cancer patients receiving care at Nganda Hospital Center in Kinshasa, DRC. Nganda Hospital is the only facility in the country equipped with a radiotherapy unit and serves as the national referral centre for oncology care. Eligible participants were adults aged ≥18 years with a histopathologically or clinically confirmed cancer diagnosis who were actively receiving care at the study site. Individuals were excluded if they declined participation or were unable to provide written informed consent. Between February and December 2025, a total of 460 patients were enrolled. The study was approved by the Ethics Committee of Kinshasa School of Public Health and the Albert Einstein College of Medicine Institutional Review Board. All participants provided written informed consent prior to enrollment.

### Data collection and measures

Trained study staff administered a structured questionnaire in the participant’s preferred language (French or Lingala) using tablets via REDCap. The questionnaire captured sociodemographic characteristics (age, sex, education, occupation, marital status, and others); healthcare access factors (distance from hospital, means of transport, transport cost, out-of-pocket medical expenses); lifestyle factors (smoking status, alcohol use); comorbidities (HIV status, hepatitis B infection, history of anogenital warts, family history of cancer); and cancer-related stigma.

Cancer-related stigma was assessed using the Cho et al. 3-factor validated scale, which measures three constructs: (a) Impossibility of recovery: perceived incurability of cancer; (b) Stereotypes: negative societal attitudes toward people with cancer; and (c) Social discrimination: perceived or experienced exclusion of people with cancer (13). Each subscale was categorized into tertiles (Low [T1], Medium [T2], High [T3]) based on the observed distribution in this sample. Data on cancer diagnosis, cancer site (coded according to ICD-10), and HIV status were abstracted from medical records at Nganda Hospital.

The socioeconomic status (SES) was constructed using principal components analysis (PCA) of 13 household-level variables, following established methods (14,15). This included ten durable household asset and utility ownership variables: electricity access, separate kitchen ownership, radio ownership, television ownership, mobile phone ownership, refrigerator ownership, bicycle ownership, motorcycle ownership, car ownership, and cooking fuel type (categorized as wood/charcoal vs. gas/electric stove). Additionally, three housing characteristic variables were included: number of rooms, number of beds, and household crowding (calculated as the ratio of household members to number of rooms or number of beds). The first principal component, which captured the largest proportion of variance (23.4%) across these indicators and represented the underlying socioeconomic gradient, was retained as the wealth index score. The resulting continuous score was categorized into tertiles with higher values indicating higher socioeconomic status. Out-of-pocket expenses were defined as the proportion of the patient’s income spent on cancer-related costs, dichotomized at the 50% threshold.

### Outcome variables

We calculated the self-reported time (in months) from the first reported symptom to cancer diagnosis, and from cancer diagnosis to treatment initiation. We then examined two primary outcomes: 1) delay in diagnosis and 2) delay in treatment. Consistent with prior literature, delays were defined using clinically meaningful thresholds: prolonged diagnostic delay was defined as >6 months, and treatment delay as >3 months, reflecting evidence that delays of this magnitude are associated with adverse cancer outcomes (7,16).

### Statistical Analysis

Continuous variables were summarized as mean (standard deviation [SD]) or median (interquartile range [IQR]), depending on the normality of the distribution, and as frequency (percentage) for categorical variables, stratified by delay group. Delay times (in months) were right-skewed; therefore, we used log-normal regression models, fitting ordinary least-squares models to the log-transformed outcomes and expressing results as geometric mean ratios (GMRs), which represent the multiplicative effect on the geometric mean delay.

Univariable log-normal models were fitted for each covariate and outcome. Variables with a p-value <0.20 in univariable analyses were considered for inclusion in multivariable models. Variable selection was performed separately for each outcome, resulting in outcome-specific multivariable models. Age, sex, and socioeconomic status (SES), measured as wealth index tertiles, were included a priori in all multivariable models regardless of statistical significance, given their established clinical relevance and potential for confounding. Results are presented as adjusted geometric mean ratios (aGMRs) with 95% confidence intervals (CIs) and two-sided p-values. All statistical tests were two-sided, with a significance level of α = 0.05. Statistical analyses were performed using R (version 4.5.2).

## RESULTS

### Participants and clinical characteristics

A total of 460 cancer patients were included in the diagnostic delay analysis, of whom 312 had treatment initiation data. The largest age group was 50-64 years (42.0%), and most participants were women (76.2%) and married (64.8%). Although over 90% had at least secondary education, 74.0% were not recently employed. Public transport was the main mode of travel (55.9%), 46.5% lived >10 km from the hospital, and 42.6% reported out-of-pocket health expenses accounting for ≥50% of household health costs. Alcohol use (12.6%) and smoking (3.1%) were uncommon. Full sociodemographic and access characteristics are presented in Table 1.

**Table 1.** Sociodemographic, clinical, and cancer care characteristics of the study population.

| Characteristic |  | Delay in diagnosis |  | Delay in treatment |  |
| --- | --- | --- | --- | --- | --- |
|  | All participants | ≤6 months | >6 months | ≤3 months | >3 months |
| **Total observations** | 460 | 207 | 253 | 158 | 154 |
| **DEMOGRAPHIC FACTORS** |  |  |  |  |  |
| **Age group (years), n (%)** |  |  |  |  |  |
| <50 | 157 (34.2) | 77 (37.2) | 80 (31.7) | 65 (41.4) | 48 (31.2) |
| 50-64 | 193 (42) | 85 (41.1) | 108 (42.9) | 57 (36.3) | 67 (43.5) |
| ≥65 | 109 (23.7) | 45 (21.7) | 64 (25.4) | 35 (22.3) | 39 (25.3) |
| **Sex, n (%)** |  |  |  |  |  |
| Men | 109 (23.8) | 37 (17.9) | 72 (28.7) | 36 (22.8) | 40 (26) |
| Women | 349 (76.2) | 170 (82.1) | 179 (71.3) | 122 (77.2) | 114 (74) |
| **Marital status, n (%)** |  |  |  |  |  |
| Single | 162 (35.2) | 75 (36.2) | 87 (34.4) | 53 (33.5) | 54 (35.1) |
| Married | 298 (64.8) | 132 (63.8) | 166 (65.6) | 105 (66.5) | 100 (64.9) |
| **Education level, n (%)** |  |  |  |  |  |
| Primary/Secondary | 266 (58.5) | 119 (58) | 147 (58.8) | 85 (53.8) | 83 (55) |
| Tertiary | 189 (41.5) | 86 (42) | 103 (41.2) | 73 (46.2) | 68 (45) |
| **Occupation, n (%)** |  |  |  |  |  |
| Employee | 70 (15.3) | 42 (20.3) | 28 (11.2) | 32 (20.3) | 18 (11.8) |
| Trader/seller | 49 (10.7) | 20 (9.7) | 29 (11.6) | 18 (11.4) | 15 (9.8) |
| Has not worked recently | 339 (74) | 145 (70) | 194 (77.3) | 108 (68.4) | 120 (78.4) |
| **SOCIOECONOMIC & HEALTHCARE ACCESS** |  |  |  |  |  |
| **SES in tertile, n (%)** |  |  |  |  |  |
| T1 (Low) | 155 (33.7) | 66 (31.9) | 89 (35.2) | 53 (33.5) | 66 (42.9) |
| T2 (Middle) | 151 (32.8) | 71 (34.3) | 80 (31.6) | 57 (36.1) | 46 (29.9) |
| T3 (High) | 154 (33.5) | 70 (33.8) | 84 (33.2) | 48 (30.4) | 42 (27.3) |
| **Transport cost (USD), n (%)** |  |  |  |  |  |
| ≤10 | 289 (62.8) | 132 (63.8) | 157 (62.1) | 101 (63.9) | 103 (66.9) |
| >10 | 171 (37.2) | 75 (36.2) | 96 (37.9) | 57 (36.1) | 51 (33.1) |
| **Distance from hospital, n (%)** |  |  |  |  |  |
| <10 km | 246 (53.5) | 115 (55.6) | 131 (51.8) | 89 (56.3) | 77 (50) |
| ≥10 km | 214 (46.5) | 92 (44.4) | 122 (48.2) | 69 (43.7) | 77 (50) |
| **Means of transport, n (%)** |  |  |  |  |  |
| Personal vehicle | 96 (20.9) | 50 (24.2) | 46 (18.2) | 34 (21.5) | 31 (20.1) |
| Public transport | 257 (55.9) | 126 (60.9) | 131 (51.8) | 89 (56.3) | 87 (56.5) |
| Taxi/Motorbike | 107 (23.3) | 31 (15) | 76 (30) | 35 (22.2) | 36 (23.4) |
| **Out-of-pocket expenses, n (%)** |  |  |  |  |  |
| <50% | 256 (57.4) | 123 (61.5) | 133 (54.1) | 84 (54.9) | 86 (58.1) |
| ≥50% | 190 (42.6) | 77 (38.5) | 113 (45.9) | 69 (45.1) | 62 (41.9) |
| **DIAGNOSTIC & TREATMENT DELAYS** |  |  |  |  |  |
| **Time from symptoms onset to diagnosis (months)** |  |  |  |  |  |
| Median (IQR) | 7.1 (2.9-17.8) | - | - | - | - |
| Mean ± SD | 15.6 +/- 26.3 | - | - | - | - |
| **Time from diagnosis to treatment initiation (months)** |  |  |  |  |  |
| Median (IQR) | 2.9 (1.2-7.1) | - | - | - | - |
| Mean ± SD | 7.4 +/- 12.8 | - | - | - | - |
| **CLINICAL & BEHAVIOURAL FACTORS** |  |  |  |  |  |
| **Alcohol consumption, n (%)** |  |  |  |  |  |
| No | 401 (87.4) | 175 (84.5) | 226 (89.7) | 142 (89.9) | 128 (83.7) |
|  | All participants | ≤6 months | >6 months | ≤3 months | >3 months |
| Yes | 58 (12.6) | 32 (15.5) | 26 (10.3) | 16 (10.1) | 25 (16.3) |
| **Smoking status, n (%)** |  |  |  |  |  |
| Ever smoked | 14 (3.1) | 2 (1) | 12 (4.8) | 3 (1.9) | 5 (3.3) |
| Living with HIV, n (%) | 15 (3.3) | 9 (4.3) | 6 (2.4) | 3 (1.9) | 5 (3.2) |
| Hepatitis B, n (%) | 14 (3.1) | 4 (1.9) | 10 (4) | 2 (1.3) | 10 (6.5) |
| History of warts, n (%) | 20 (4.4) | 11 (5.3) | 9 (3.6) | 4 (2.6) | 6 (3.9) |
| Family history of cancer, n (%) | 125 (27.2) | 58 (28) | 67 (26.5) | 45 (28.5) | 46 (29.9) |
| **Current treatment regimen, n (%)** |  |  |  |  |  |
| Has not yet started treatment | 3 (1.0) | - | - | 2 (1.3) | 1 (0.7) |
| Chemotherapy | 124 (40.7) | - | - | 74 (48.1) | 50 (33.1) |
| Radiotherapy | 98 (32.1) | - | - | 47 (30.5) | 51 (33.8) |
| Surgery | 2 (0.7) | - | - | 1 (0.6) | 1 (0.7) |
| Follow-up/Surveillance | 78 (25.6) | - | - | 30 (19.5) | 48 (31.8) |
| **PERCEIVED CANCER-RELATED STIGMA** |  |  |  |  |  |
| **(a) Impossibility of Recovery, n (%)** |  |  |  |  |  |
| Low (T1) | 154 (33.6) | 79 (38.2) | 75 (29.8) | 51 (32.3) | 44 (28.8) |
| Medium (T2) | 153 (33.3) | 67 (32.4) | 86 (34.1) | 49 (31) | 56 (36.6) |
| High (T3) | 152 (33.1) | 61 (29.5) | 91 (36.1) | 58 (36.7) | 53 (34.6) |
| **(b) Stereotypes, n (%)** |  |  |  |  |  |
| Low (T1) | 153 (33.3) | 76 (36.7) | 77 (30.6) | 51 (32.3) | 41 (26.8) |
| Medium (T2) | 154 (33.6) | 70 (33.8) | 84 (33.3) | 66 (41.8) | 47 (30.7) |
| High (T3) | 152 (33.1) | 61 (29.5) | 91 (36.1) | 41 (25.9) | 65 (42.5) |
| **(c) Social Discrimination, n (%)** |  |  |  |  |  |
| Low (T1) | 154 (33.6) | 65 (31.4) | 89 (35.3) | 43 (27.2) | 50 (32.7) |
| Medium (T2) | 153 (33.3) | 84 (40.6) | 69 (27.4) | 74 (46.8) | 43 (28.1) |
| High (T3) | 152 (33.1) | 58 (28) | 94 (37.3) | 41 (25.9) | 60 (39.2) |

1. SES = socioeconomic status; IQR = interquartile range; SD = standard deviation. 2. Socioeconomic status quintiles were derived from a wealth index based on household assets. 3. Cancer stigma was measured using the 12-item scale developed by Cho et al. (2013). 4. Outcome variables were categorized to reflect clinically meaningful delay thresholds cutoffs. Time from symptom onset to diagnosis was dichotomized as ≤6 months vs. >6 months. Time from diagnosis to treatment initiation was dichotomized as ≤3 months vs. >3 months.

HIV co-infection was present in 15 participants (3.3%), hepatitis B in 14 (3.1%), and a history of anogenital warts in 20 (4.4%). A family history of cancer was reported in 27.2% of participants. Among the 312 patients in the treatment delay cohort, chemotherapy was the most commonly received treatment modality (48.1% among those with short delays; 33.1% among those with longer delays), followed by radiotherapy (30.5% and 33.8%, respectively).

### Delays in the cancer care cascade

Prolonged delays were observed at both stages of the cancer care cascade. The median time from symptom onset to diagnosis was 7.1 months (IQR: 2.9-17.8), with 55.0% of participants experiencing delays >6 months. The median time from diagnosis to treatment initiation was 2.9 months (IQR: 1.2-7.1), and 49.4% experienced delays >3 months. Compared with participants without prolonged treatment delay, those with prolonged delay were less likely to receive chemotherapy (33.1% vs. 48.1%) and more likely to be under follow-up or surveillance (31.8% vs. 19.5%), while radiotherapy use was similar between groups (Table 1).

### Cancer site distribution

The distribution of cancer diagnoses differed markedly by sex, with the overall pattern underscoring a substantial burden of malignancies related to the reproductive system (Figure 1). Among women, breast cancer was the most prevalent malignancy (52.6%), followed by cervical cancer (33.3%) and endometrial/uterine cancer (1.2%), together accounting for more than 87% of female diagnoses. Among men, prostate cancer (32.7%) and head and neck cancers (26.2%) were the most common diagnoses, with colorectal cancer accounting for an additional 7.5%. Liver cancer, peripheral and musculoskeletal tumors, central nervous system malignancies, and lung or tracheal cancers each accounted for a small proportion of cases in both sexes.

**Figure 1.**
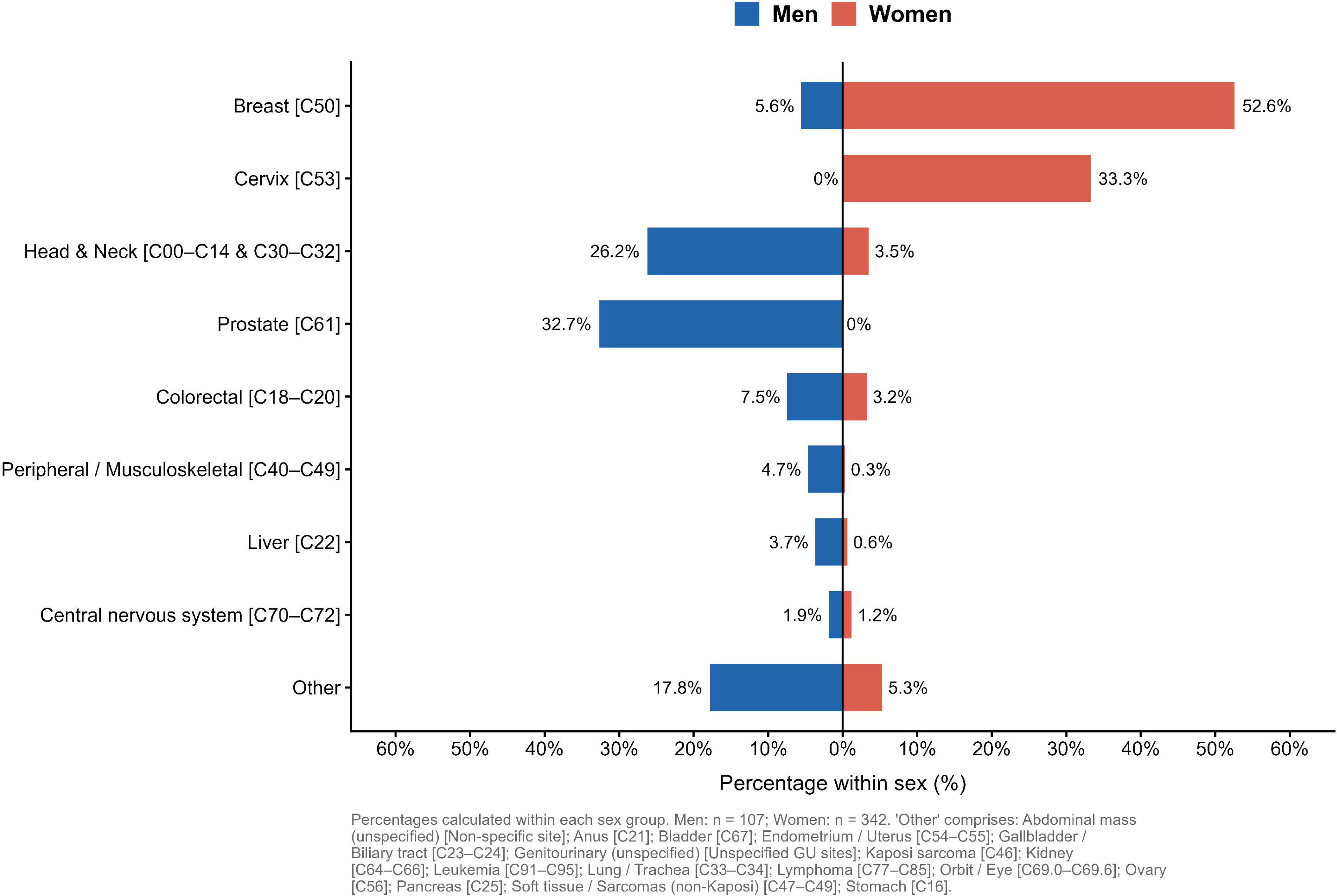

### Factors associated with delayed diagnosis

Full univariable associations are presented in Table 2. After multivariable adjustment, six factors were independently associated with delayed diagnosis (Figure 2). Older age was associated with longer time to diagnosis: compared with patients aged <50 years, those aged ≥65 years had a 55% longer time to diagnosis (aGMR 1.55, 95% CI: 1.03-2.31). Occupational status was a strong predictor of diagnostic delay: compared with formally employed patients, those engaged in trading or selling had nearly twice the time to diagnosis (aGMR 1.99, 95% CI: 1.21-3.28), while those not working recently experienced a 68% longer delay (aGMR 1.68, 95% CI: 1.15-2.47). Among transport modalities, reliance on taxi or motorbike was linked to a 92% longer diagnostic delay relative to use of a personal vehicle (aGMR 1.92, 95% CI: 1.25-2.94). A history of smoking was associated with more than a twofold delay (aGMR 2.25, 95% CI: 1.03-4.91), and hepatitis B co-infection was also associated with longer delay (aGMR 1.88, 95% CI: 1.06-3.34). Finally, medium levels of perceived social discrimination stigma were associated with a 33% shorter time to diagnosis compared with low levels (aGMR 0.67, 95% CI: 0.48-0.93).

**Figure 2.**
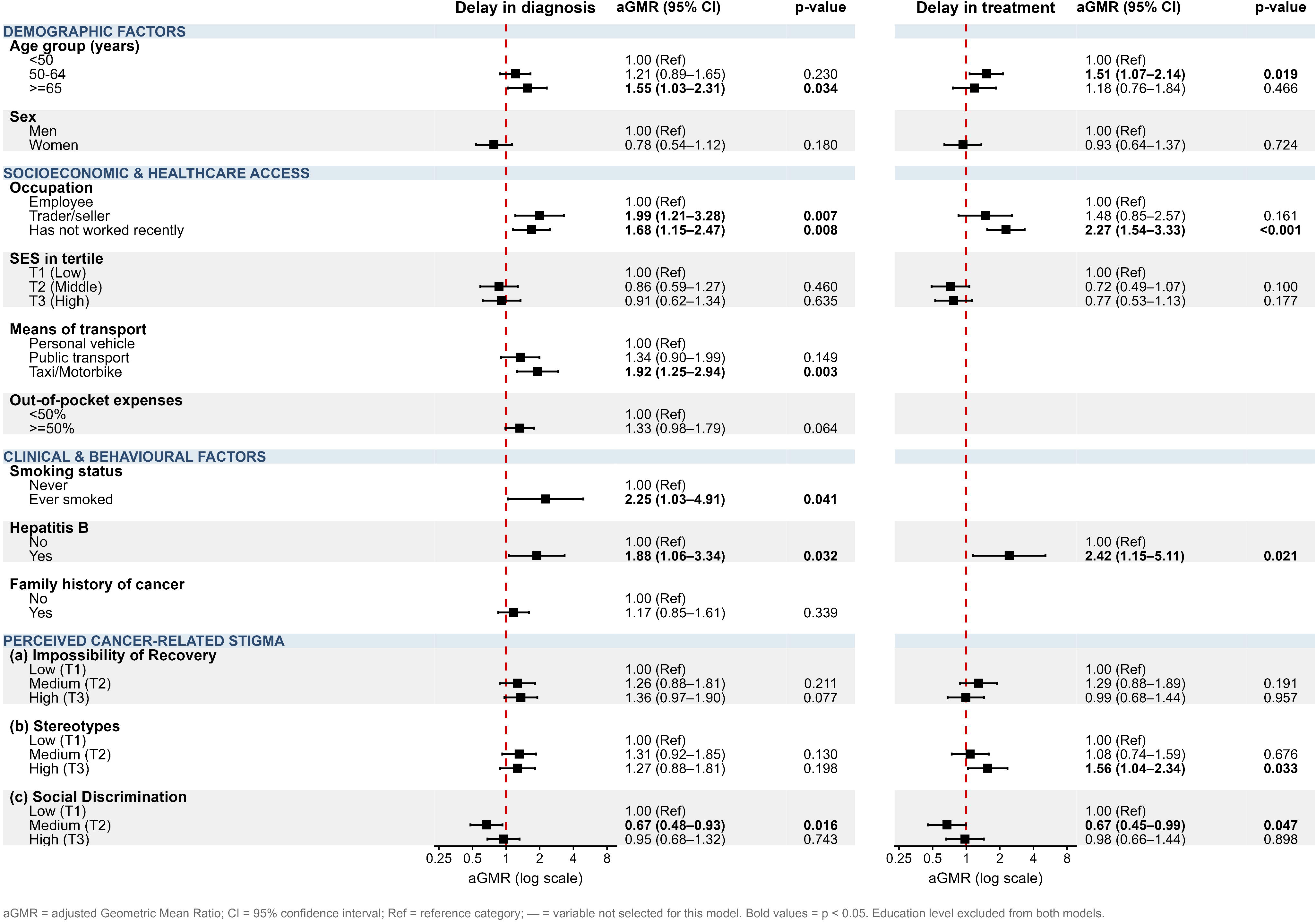

**Table 2.** Univariate analysis of demographic, socioeconomic, clinical, and stigma-related factors associated with delay in care.

|  | Delay in diagnosis<br>(log-normal, continuous) |  |  | Delay in treatment<br>(log-normal, continuous) |  |  |
| --- | --- | --- | --- | --- | --- | --- |
| Characteristic | GMR | 95% CI | p-value | GMR | 95% CI | p-value |
| **DEMOGRAPHIC FACTORS** |  |  |  |  |  |  |
| **Age group (years)** |  |  |  |  |  |  |
| <50 | 1.00 | Ref |  | 1.00 | Ref |  |
| 50-64 | 1.15 | (0.86–1.53) | 0.362 | 1.56 | (1.10–2.22) | 0.012 |
| >=65 | 1.58 | (1.09–2.27) | 0.015 | 1.30 | (0.86–1.98) | 0.215 |
| **Sex** |  |  |  |  |  |  |
| Men | 1.00 | Ref |  | 1.00 | Ref |  |
| Women | 0.67 | (0.48–0.93) | 0.018 | 0.87 | (0.61–1.23) | 0.420 |
| **Marital status** |  |  |  |  |  |  |
| Married | 1.00 | Ref |  | 1.00 | Ref |  |
| Single | 0.98 | (0.75–1.28) | 0.888 | 0.91 | (0.66–1.26) | 0.560 |
| **SOCIOECONOMIC & HEALTHCARE ACCESS** |  |  |  |  |  |  |
| **Education level** |  |  |  |  |  |  |
| Primary/Secondary | 1.00 | Ref |  | 1.00 | Ref |  |
| Tertiary | 1.01 | (0.77–1.31) | 0.952 | 0.93 | (0.68–1.26) | 0.624 |
| **Occupation** |  |  |  |  |  |  |
| Employee | 1.00 | Ref |  | 1.00 | Ref |  |
| Trader/seller | 1.53 | (0.96–2.44) | 0.071 | 1.27 | (0.72–2.23) | 0.405 |
| Has not worked recently | 1.48 | (1.05–2.07) | 0.024 | 2.25 | (1.55–3.27) | <0.001 |
| **SES in tertile** |  |  |  |  |  |  |
| T1 (Low) | 1.00 | Ref |  | 1.00 | Ref |  |
| T2 (Middle) | 0.90 | (0.64–1.27) | 0.547 | 0.82 | (0.56–1.19) | 0.290 |
| T3 (High) | 0.99 | (0.72–1.35) | 0.937 | 0.77 | (0.54–1.11) | 0.167 |
| **Means of transport** |  |  |  |  |  |  |
| Personal vehicle | 1.00 | Ref |  | 1.00 | Ref |  |
| Public transport | 1.08 | (0.77–1.50) | 0.665 | 0.93 | (0.62–1.39) | 0.715 |
| Taxi/Motorbike | 1.77 | (1.22–2.57) | 0.003 | 0.93 | (0.58–1.51) | 0.779 |
| **Out-of-pocket expenses** |  |  |  |  |  |  |
| <50% | 1.00 | Ref |  | 1.00 | Ref |  |
| >=50% | 1.25 | (0.95–1.63) | 0.112 | 0.84 | (0.61–1.15) | 0.277 |
| **Distance from hospital** |  |  |  |  |  |  |
| <10 km | 1.00 | Ref |  | 1.00 | Ref |  |
| >=10 km | 1.09 | (0.84–1.41) | 0.524 | 1.05 | (0.77–1.43) | 0.778 |
| **Transport cost (USD)** |  |  |  |  |  |  |
| <=10 | 1.00 | Ref |  | 1.00 | Ref |  |
| >10 | 1.05 | (0.80–1.38) | 0.706 | 1.06 | (0.77–1.47) | 0.706 |
| **CLINICAL & BEHAVIOURAL FACTORS** |  |  |  |  |  |  |
| **Smoking status** |  |  |  |  |  |  |
| Never | 1.00 | Ref |  | 1.00 | Ref |  |
| Ever smoked | 2.15 | (1.06–4.39) | 0.035 | 1.18 | (0.53–2.64) | 0.679 |
| **Alcohol consumption** |  |  |  |  |  |  |
| No | 1.00 | Ref |  | 1.00 | Ref |  |
| Yes | 0.96 | (0.65–1.43) | 0.845 | 0.89 | (0.52–1.52) | 0.667 |
| Characteristic | GMR | 95% CI | p-value | GMR | 95% CI | p-value |
| **Hepatitis B** |  |  |  |  |  |  |
| No | 1.00 | Ref |  | 1.00 | Ref |  |
| Yes | 2.12 | (1.17–3.83) | 0.013 | 3.02 | (1.51–6.04) | 0.002 |
| **Family history of cancer** |  |  |  |  |  |  |
| No | 1.00 | Ref |  | 1.00 | Ref |  |
| Yes | 1.24 | (0.91–1.67) | 0.168 | 0.89 | (0.65–1.22) | 0.462 |
| **Living with HIV** |  |  |  |  |  |  |
| No | 1.00 | Ref |  | 1.00 | Ref |  |
| Yes | 0.66 | (0.26–1.69) | 0.389 | 0.84 | (0.46–1.55) | 0.578 |
| **History of warts** |  |  |  |  |  |  |
| No | 1.00 | Ref |  | 1.00 | Ref |  |
| Yes | 0.71 | (0.30–1.68) | 0.438 | 1.39 | (0.56–3.41) | 0.476 |
| **PERCEIVED CANCER-RELATED STIGMA** |  |  |  |  |  |  |
| **(a) Impossibility of Recovery** |  |  |  |  |  |  |
| Low (T1) | 1.00 | Ref |  | 1.00 | Ref |  |
| Medium (T2) | 1.29 | (0.94–1.78) | 0.119 | 1.42 | (0.96–2.08) | 0.075 |
| High (T3) | 1.35 | (0.99–1.84) | 0.061 | 1.18 | (0.80–1.72) | 0.405 |
| **(b) Stereotypes** |  |  |  |  |  |  |
| Low (T1) | 1.00 | Ref |  | 1.00 | Ref |  |
| Medium (T2) | 1.20 | (0.86–1.67) | 0.279 | 0.94 | (0.63–1.39) | 0.753 |
| High (T3) | 1.41 | (1.01–1.97) | 0.041 | 1.59 | (1.06–2.40) | 0.025 |
| **(c) Social Discrimination** |  |  |  |  |  |  |
| Low (T1) | 1.00 | Ref |  | 1.00 | Ref |  |
| Medium (T2) | 0.71 | (0.52–0.97) | 0.032 | 0.73 | (0.50–1.07) | 0.105 |
| High (T3) | 1.11 | (0.81–1.53) | 0.510 | 1.19 | (0.81–1.75) | 0.370 |

Univariate log-normal regression models fitted to log-transformed delay times (months). Results expressed as Geometric Mean Ratios (GMR) with 95% confidence intervals

Two variables that were significantly associated with diagnostic delay in univariable analyses were attenuated to non-significance after multivariable adjustment, suggesting potential confounding by other covariates; however, limited statistical power cannot be excluded. Female sex was associated with a relative 67% shorter time to diagnosis (GMR 0.67, 95% CI: 0.48-0.93), whereas high levels of cancer-related stereotype stigma were associated with a relative 41% longer time to diagnosis (GMR 1.41, 95% CI: 1.01-1.97); neither association was retained after adjustment.

### Factors associated with delayed treatment initiation

Age 50-64 years was associated with a relative 51% longer time to treatment initiation compared with patients aged <50 years (aGMR 1.51, 95% CI: 1.07-2.14). Not being currently employed was the strongest socioeconomic predictor, with these patients experiencing more than twice the time to treatment initiation compared with those in formal employment (aGMR 2.27, 95% CI: 1.54-3.33), indicating a consistent barrier across the cancer care pathway. Hepatitis B co-infection was also a strong and independent predictor, associated with a similarly substantial increase in time to treatment initiation (aGMR 2.42, 95% CI: 1.15-5.11), highlighting a potentially important clinical vulnerability within the care pathway.

Cancer-related stereotype stigma was independently associated with longer time to treatment initiation, with high stigma levels associated with a relative 56% longer delay compared with low levels (aGMR 1.56, 95% CI: 1.04-2.34). Perceived social discrimination at moderate levels was also independently associated with shorter treatment delays (aGMR 0.67, 95% CI: 0.45-0.99; p = 0.047).

## DISCUSSION

In this study, more than half of patients had a symptom-to-diagnosis interval exceeding 6 months, and approximately half had a diagnosis-to-treatment interval exceeding 3 months, findings that are consistent with patterns reported across SSA (6,17). These intervals are substantially longer than those reported in high-income settings, where diagnostic intervals are typically measured in weeks to a few months and treatment is often initiated within 30-60 days (18,19).

Multivariable analyses identified several determinants of delay, some of which were consistent across both diagnostic and treatment delays, while others differed between the two phases. Not being currently employed emerged as a consistent predictor of both diagnostic and treatment delays, underscoring the central role of socioeconomic vulnerability. Hepatitis B co-infection was also associated with longer delays at both stages of care, suggesting an important clinical subgroup at risk of delayed care.

Transport-related barriers and smoking were primarily associated with delayed diagnosis, while older age (50-64 years) and cancer-related stereotype stigma were associated with longer delays in treatment initiation. Moderate perceived social discrimination was associated with shorter time to diagnosis, with a statistically significant association also observed for treatment. These findings highlight the combined influence of structural, clinical, and psychosocial factors on timely access to cancer care in this setting.

Among these determinants, unemployment emerged as the most consistent predictor of longer delays in both diagnosis and treatment initiation. Notably, 74% of participants reported not working recently, compared with an estimated 35% of working-age adults in the general population outside the labor force (20), suggesting substantial socioeconomic vulnerability. Informal work, such as trading or selling, was also associated with longer diagnostic delay, highlighting the vulnerability of individuals lacking employment-related protections and facing income loss during illness. Despite CNLC subsidies for chemotherapy and radiotherapy, residual out-of-pocket costs for diagnostics, transport, and supportive care likely continue to delay timely cancer care among economically disadvantaged patients.

Hepatitis B co-infection was associated with longer delays in both diagnosis and treatment initiation. Given the endemicity of hepatitis B in the DRC and across SSA (21,22), this association may reflect fragmented care pathways, increased clinical complexity related to underlying liver disease, and overlapping symptoms that delay cancer investigation and treatment. Hepatitis B may also act as a marker of broader socioeconomic and health system vulnerabilities, highlighting the need for more integrated hepatology-oncology care pathways.

The association between reliance on taxi or motorbike transport and longer time to diagnosis likely reflects underlying socioeconomic and geographic barriers to care. Use of informal transport may indicate limited financial resources and residence in peri-urban or underserved areas with restricted access to formal public transport. Similar patterns have been reported across SSA, where transport-related costs, distance, and health system barriers are major determinants of delayed cancer care (23–25). These findings support the need for decentralization of diagnostic services and financial subsidies for transport to secondary and tertiary care facilities, approaches being piloted in several SSA countries (24,25).

A history of smoking was associated with more than a twofold increase in time to diagnosis. Although smoking prevalence was low in this predominantly female sample, consistent with the low prevalence of tobacco use among women in the DRC (26), this association may indicate that smoking acts as a marker of broader vulnerability or delayed care-seeking behaviors. Smoking may also be correlated with other unmeasured socioeconomic or behavioral factors that influence healthcare access, including lower engagement with preventive services. Given the relatively small number of smokers in this study population, this finding should be interpreted with caution and warrants confirmation in larger studies.

One of the most novel findings was the independent association between cancer-related stigma and delayed treatment initiation. Higher stereotype-related stigma was associated with longer time to treatment, independent of socioeconomic and clinical factors, possibly because internalized stigma leads patients to conceal their diagnosis or avoid care. Although such dynamics have been described qualitatively in SSA (27,28), quantitative evidence remains limited. The inverse association between moderate perceived social discrimination and shorter delays was unexpected and warrants further investigation and will be explored in ongoing qualitative and longitudinal analyses.

Older age was independently associated with longer diagnostic delays, consistent with evidence suggesting that older individuals may present later, potentially due to normalization of symptoms, mobility limitations, or age-related perceptions of illness (29). Targeted awareness strategies focusing on older adults may help reduce this delay. Notably, variables commonly hypothesized to impede access to care, such as lower SES, greater distance from the hospital, and higher out-of-pocket costs, were not independently associated with delays. This may indicate that employment status more accurately captures the economically relevant dimension of access in this setting than asset-based wealth measures. Finally, despite being a key a priori hypothesis, HIV status was not associated with delay in cancer care in either univariable or multivariable analyses, likely reflecting limited statistical power given the small number of participants living with HIV enrolled in this study (n = 15; 3.3% of the total sample).

These findings should be interpreted in light of limitations. The cross-sectional design precludes causal inference, and reliance on self-reported symptom onset may have introduced recall bias. Additionally, the possibility of reverse causality cannot be ruled out; for instance, a cancer diagnosis and its subsequent treatment burden may lead to unemployment, rather than pre-existing unemployment serving as the primary driver of diagnostic delay. This single-center study may also limit generalizability and underestimate the true burden of delays, as patients unable to reach Nganda Hospital Center, particularly those outside Kinshasa, were not captured.

Finally, while the overall sample size was adequate for primary analyses, the limited number of participants in certain subgroups, particularly people living with HIV, may have reduced statistical power to detect meaningful associations.

In conclusion, substantial delays were observed along the cancer care pathway in this setting, reflecting a complex interplay of socioeconomic vulnerability, clinical comorbidity, transport-related barriers, and cancer-related stigma. These findings suggest that delays are not solely driven by structural constraints but also by behavioral and psychosocial factors that influence care-seeking and treatment initiation.

Addressing these delays will require integrated strategies, including strengthening financial protection, improving coordination of care for patients with comorbid conditions, decentralizing diagnostic services, and implementing interventions to reduce cancer-related stigma. With ongoing follow-up, longitudinal analyses of this cohort will help confirm these findings and clarify underlying mechanisms in similar settings.

## Data Availability

De-identified individual participant data may be made available to qualified researchers upon reasonable request to the corresponding author and after approval by the Kinshasa School of Public Health Ethics Committee and the Albert Einstein College of Medicine IRB.

## Author Contributions

Conception and design: JCD, NZ, RK, MY. Data collection and curation: KS, ASA, FLK, IY. Statistical analysis: JCD, NZ. Interpretation of results: all authors. Drafting of the manuscript: JCD. Critical revision of the manuscript for important intellectual content: all authors. Funding acquisition and supervision: NZ, RK, MY. All authors approved the final version.

## Acknowledgments

The authors gratefully acknowledge the Nganda Team: Vandrome Nakundi, Mah-soune Apithy, Guilaine Mopoh, Abou Dao, Arnold Sulu, and Stanislas Sulu, for their valuable contributions to study planning, data collection, study conduct, and administration. We thank Nganda Hospital Center for hosting and supporting this research, and we are deeply grateful to all participants who took part in the study.

